# Race and Ethnic Group Dependent Space Radiation Cancer Risk Predictions

**DOI:** 10.1101/2021.09.08.21263281

**Authors:** Francis A. Cucinotta, Premkumar B. Saganti

**Affiliations:** Department of Health Physics and Diagnostic Sciences, University of Nevada, Las Vegas, NV, United States of America; Prairie View A&M University, Prairie View TX, United States of America

**Keywords:** Relative risk models, galactic cosmic rays (GCR), space radiation, cancer risk, multi-cultural risks

## Abstract

Future space missions by national space agencies and private industry, including space tourism, will include a diverse makeup of crewmembers with extensive variability in age, sex, and race or ethnic groups. The relative risk (RR) model is used to transfer epidemiology data between populations to estimate radiation risks. In the RR model cancer risk is assumed to be proportional to background cancer rates and limited by other causes of death, which are dependent on genetic, environmental and dietary factors that are population dependent. Here we apply the NSCR-2020 model to make the first predictions of age dependent space radiation cancer risks for several U.S. populations, which includes Asian-Pacific Islanders (API), Black, Hispanic (white and black), and White (non-Hispanic) populations. Results suggest that male API and Hispanic populations have the overall lowest cancer risks, while White females have the highest risk. Blacks have similar total cancer rates as Whites, however their reduced life expectancy leads to modestly lower lifetime radiation risks compared to Whites. There are diverse tissue specific cancer risk ranking across sex and race, which include sex specific organ risks, females having larger lung, stomach, and urinary-bladder radiation risks, and males having larger colon and brain risks.

## INTRODUCTION

The makeup of astronauts and cosmonauts have become more diverse since the first missions in the 1960’s trending to be more reflective of the proportion of male and female and racial or ethnic distributions in host countries [1]. In addition, the role of private space missions and space tourism suggest significant diversity in age, sex, race and ethnicity of space explorers that will participate in future space missions. In this report, we predict space radiation cancer risks for White, Black, Hispanic (black and white) and Asian-Pacific Islander (API) populations in the United States (US) using background rates for tissue specific cancer and all causes of death reported by the National Cancer Institute SEER [2,3] and U.S. Center of Disease Control and Prevention (CDC) [4], respectively.

Cancer is a multistage process often described in terms of initiation-promotion-progression concepts [5]. The Hallmarks of cancer [6] describe these steps in terms of aberrant changes in critical biological processes involved in tumor development, including self-sufficiency in growth signals, insensitivity to anti-growth signals, tissue invasion and metastasis, limitless replicative potential, sustained angiogenesis (blood vessel growth), and evasion of apoptosis (cell death). The multistage model, the role of cancer hallmarks, and the minimal latency between radiation exposure and tumor development, which is typically more than 5 years for solid cancer and 1 year for leukemia, suggests that radiation likely produces only a subset of the changes necessary for tumor development, while spontaneous processes influence the development and expression of cancer [7]. Galactic cosmic rays (GCR) and solar particle events (SPE) are efficient producers of DNA damage, while low doses of GCR produce inflammatory changes important in cancer development [8-11], however it is not clear if all the cancer hallmark processes are induced by low doses of space radiation (<1 Gy), which would be incurred on long-term space missions. These observations lend support to a relative risk (RR) model to transfer risk estimates across populations with varying background cancer rates. Evidence in support of the RR model was also provided by Storer et al. [12] whom studied the transfer of risk between 5 strains of male and female mice exposed to gamma-rays at moderate acute doses (< 2 Gy) in comparison to the results from the Life Span Study (LSS) of atomic bomb survivors. It was concluded that for most tumor types, susceptibility to radiation carcinogenesis is related to spontaneous incidence, while RR derived from mouse studies were consistent with those from the LSS for tumors of the lung, breast, liver and leukemia. Several other cancer types important in humans such as stomach, colon and brain were not considered in the results of Storer et al. [12] due to limitations in the types of tumors found in various mouse strains studied.

The most recent studies of LSS provides extensive parameterizations of the age and age at exposure dependence of tissue specific cancer incidence for males and females, and considers improved estimates of the influence of life-style factors such as the use of tobacco products on cancer risk [13-22]. We use these recent LSS findings and the NASA Space Cancer Risk model (NSCR-2020) [23-27] to make predictions of cancer risks from GCR for U.S. populations as a function of sex and age for Asian-Pacific Islanders (API), Black, Hispanic (Black and White), and White (non-Hispanic) populations. The NSCR uses quality factors (QF) influenced from particle track structure concepts and parameterized against extensive mouse tumor and surrogate cancer endpoints in cell culture studies. Probability distribution functions (PDFs) that estimate uncertainties in epidemiology data, organ exposures in space, dose-rate and radiation quality effects, including non-targeted effects (NTEs), are considered. We use the NSCR model to estimate cancer risks for annual GCR exposures and a Mars mission for several populations.

## Results and Discussion

Data from the Center of Disease Control and Prevention (CDC) [4] and the NIH Surveillance Epidemiology and End Results (SEER) [2,3] are used to estimate the life-table (probability of survival to a given age) and tissue specific cancer incidence and mortality rates for the time-period of 2014-2018. The life expectancy of males and females of different racial or ethnic groups in the U.S. are shown in **Table** 1. Life expectancy estimates for API’s was not available in the 2018 report [4], and we based the API estimate on the larger values compared to Hispanics (∼2 to 3 years) from other recent time periods [28]. **Figure 1** shows the survival probability for those alive at age 35 y for the various populations considered. **Table 2** summarizes the SEER age adjusted incidence rates for the different races and males and females [2]. Hispanics and API’s compared to Blacks and Whites have health advantages for both cancer and lifespan and similar advantages for circulatory diseases. Black and White males have the largest age adjusted cancer rates, while Black males have the lowest life expectancy of 71 years. For API’s a further division between Asian immigrants compared to Pacific Islanders suggest a health disparity with Pacific Islanders having overall lower life expectancy and larger cancer incidence [29]. Indeed, further division of Hispanic populations into racial groups or all groups into U.S. states or regions would reveal differences in life expectancy and cancer rates. The calculations described next illustrate the manifestation on radiation risks for a range of such differences on radiation risk predictions. Tissue specific differences in cancer rates are described by the SEER data base and used in the analyses described below.

**Table 1.** Life Expectancy in 2018 for several U.S. racial and ethnic groups for both males and females [4].

| **Race** | **Life Expectancy (y) Females** | **Life Expectancy (y) Males** |
| --- | --- | --- |
| **Asian Pacific-Islanders\*** | 86 | 81 |
| **Black** | 78.0 | 71.3 |
| **Hispanic (white and black)** | 84.3 | 79.1 |
| **White** | 81.1 | 76.2 |

\*Data for 2018 was not available and estimate is based on trend for other recent years.

**Figure 1.**
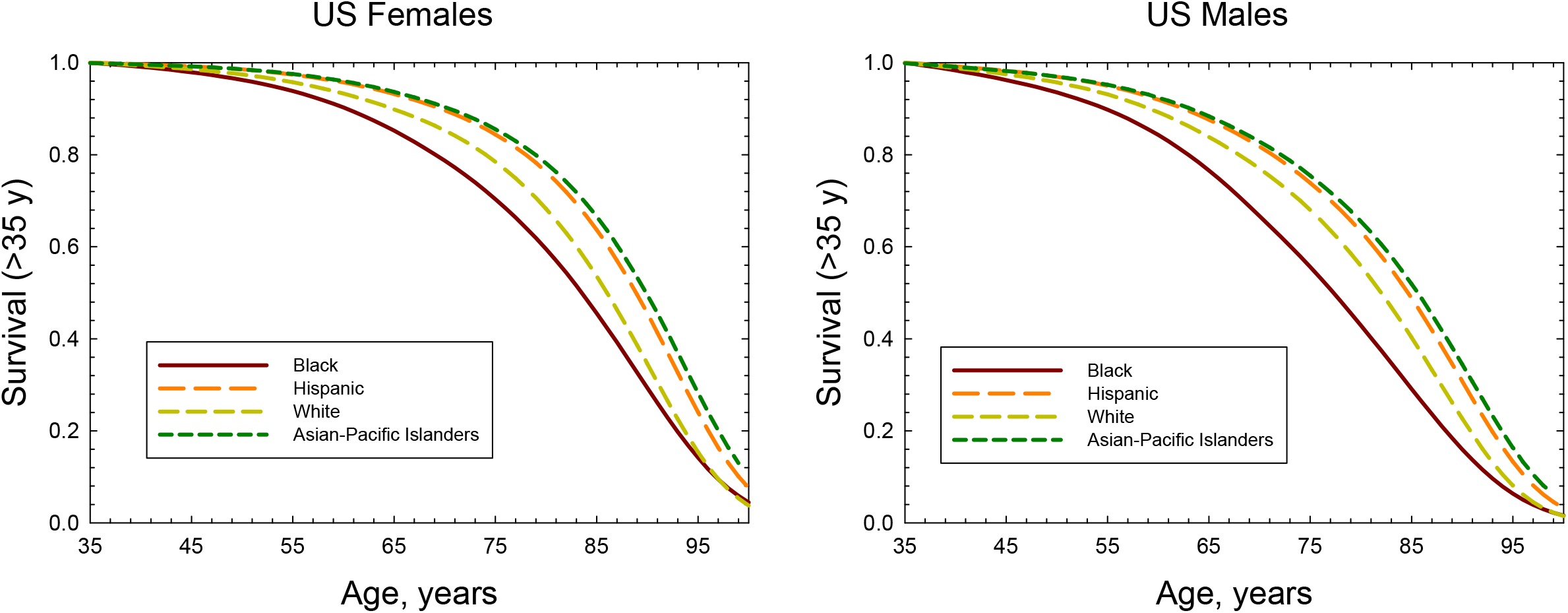
Survival probability after age 35 y for U.S. females and males of different racial or ethnic groups.

**Table 2.** Age adjusted total cancer incidence rates per 100,000 persons in SEER delay adjusted model [2,3] for 2014-2018 for several U.S. racial and ethnic groups for both males and females.

| **Race** | **Females** | **Males** |
| --- | --- | --- |
| **Asian Pacific-Islanders** | 328.2 | 311.2 |
| **Black** | 400.2 | 516.1 |
| **Hispanic (white and black)** | 355.9 | 371.0 |
| **White** | 460.2 | 520.4 |

Low LET epidemiology data is used in the NSCR model with space radiation quality factors (QF) and space radiation environmental and transport models used to make predictions for space exposures [22-24]. Results from LSS for tissue specific excess relative risk (ERR) for incidence as a function of age and latency are used as they provide data for a similar dose range as space missions and is a whole-body exposure; which is often not true of lower dose protracted occupational exposures or high dose fractionated exposures in various medical procedures. For thyroid and remainder cancers we use the BEIR VII report parametrizations of ERR [30]. We note that GCR heavy ion energy deposition in cells occurs at very high dose-rates (<10^−16^ s) with doses of 0.1 to >2 Gy deposited per cell traversed dependent on charge number and kinetic energy, which are similar doses to the acute exposures in the atomic bomb survivors. Dose-rate reduction factors for the low LET components of GCR are described using a Bayesian analysis as described previously [23].

The most recent series of LSS analyses [13-22] have longer follow-up times and improved adjustments for smoking and other lifestyle factors compared to those used in previous NSCR versions. **Table 3** show ERR parametric values (See Methods) for several tissues comparing females to males from the LSS results. Females have larger values of excess relative risk (ERR) compared to males for total solid, lung, stomach, liver, urinary-bladder cancers, and other (remainder) cancers. Males have larger ERR for colon and brain cancers. There are differences due to sex dependent cancers (breast, ovarian, and prostate), and the most recent LSS study finds a significant ERR for pancreatic cancer for females but not for males. Females and males are found to have similar ERR for leukemia (excluding chronic lymphatic leukemia (CLL)) and esophageal cancer. These age, sex and tissue specific ERR models are folded with the background rates for cancers and age dependent survival to make absolute risk predictions as described next.

**Table 3.**
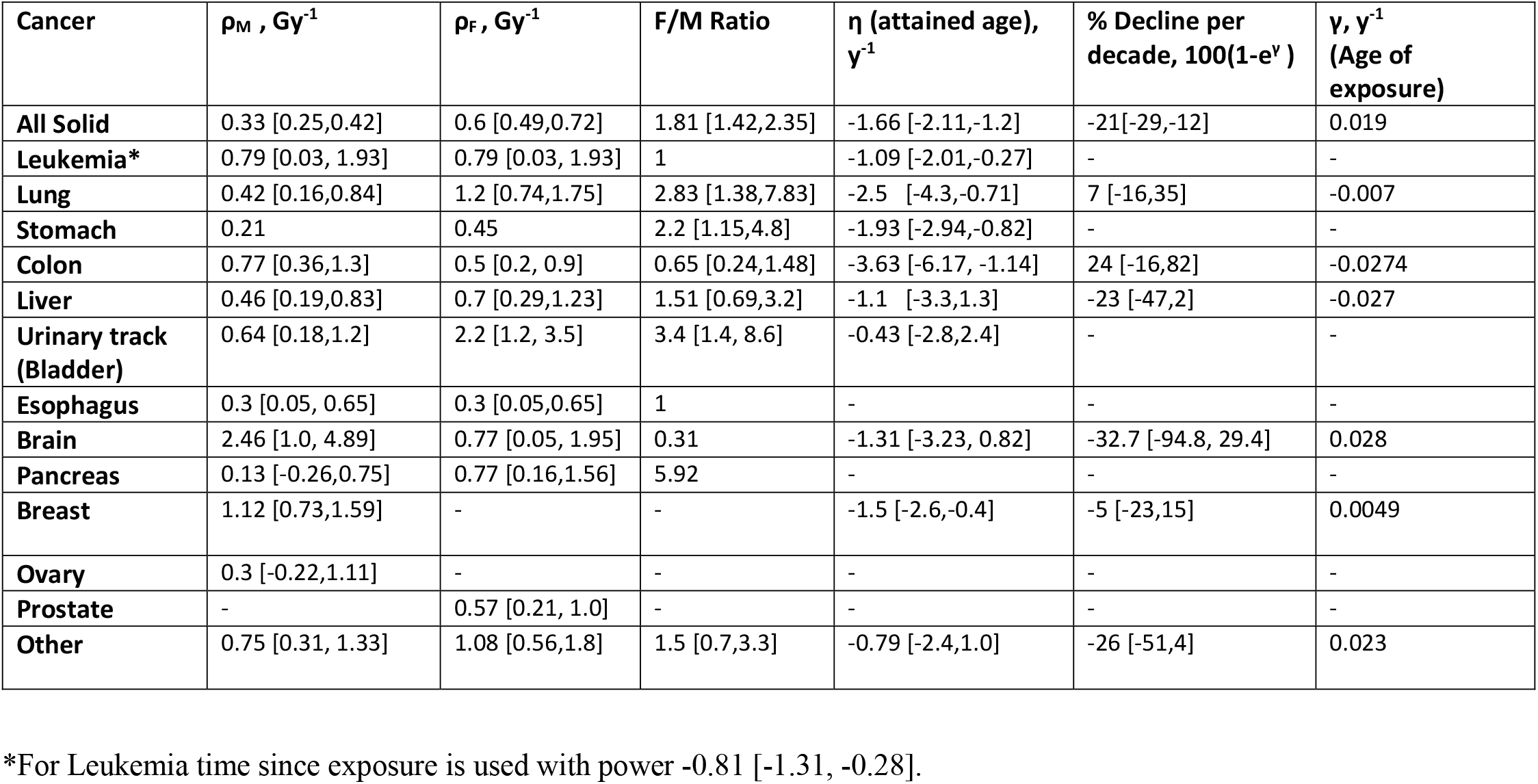
Parameters for Excess Relative Risk (ERR) models from Life Span Study (LSS) of atomic bomb survivors, and ratio of female (F) to male (M) risk for exposure at age 30 y and incidence at age 70 y. Estimates of effects of age at exposure and attained age are fit with a common model for females and males. Estimates of ERR per Sv are higher for females for most cancers with the exception of colon and brain cancers. For several tissues, values were not reported because they are non-significant. For esophagus and leukemia excluding chronic lymphatic leukemia (CLL) differences between females and males were not reported because they were essential the same. For the group of other also denoted as remainder cancers parameters from the BEIR VII report [30] are used.

We used the data described above and the NSCR model of space radiation QF’s and tissue specific particle fluence and doses to estimate the risk of exposure induced cancer (REIC) and the risk of exposure induced death (REID) for average solar minimum conditions and spacecraft shielding of 20 g/cm^2^ of aluminum. **Figure 2** shows tissue specific REIC and REID predictions for females for 1-year GCR exposures at average solar minimum conditions. White females have the largest REIC values for leukemia, urinary bladder, brain, lung, breast, remainder and total cancer amongst female groups. Hispanics and APIs have the largest REIC for stomach cancers. A similar comparison is shown in **Figure 3** for males. The differences between groups for males is smaller than females, however Black and White males have a modestly larger total REIC values than APIs or Hispanic.

**Figure 2.**
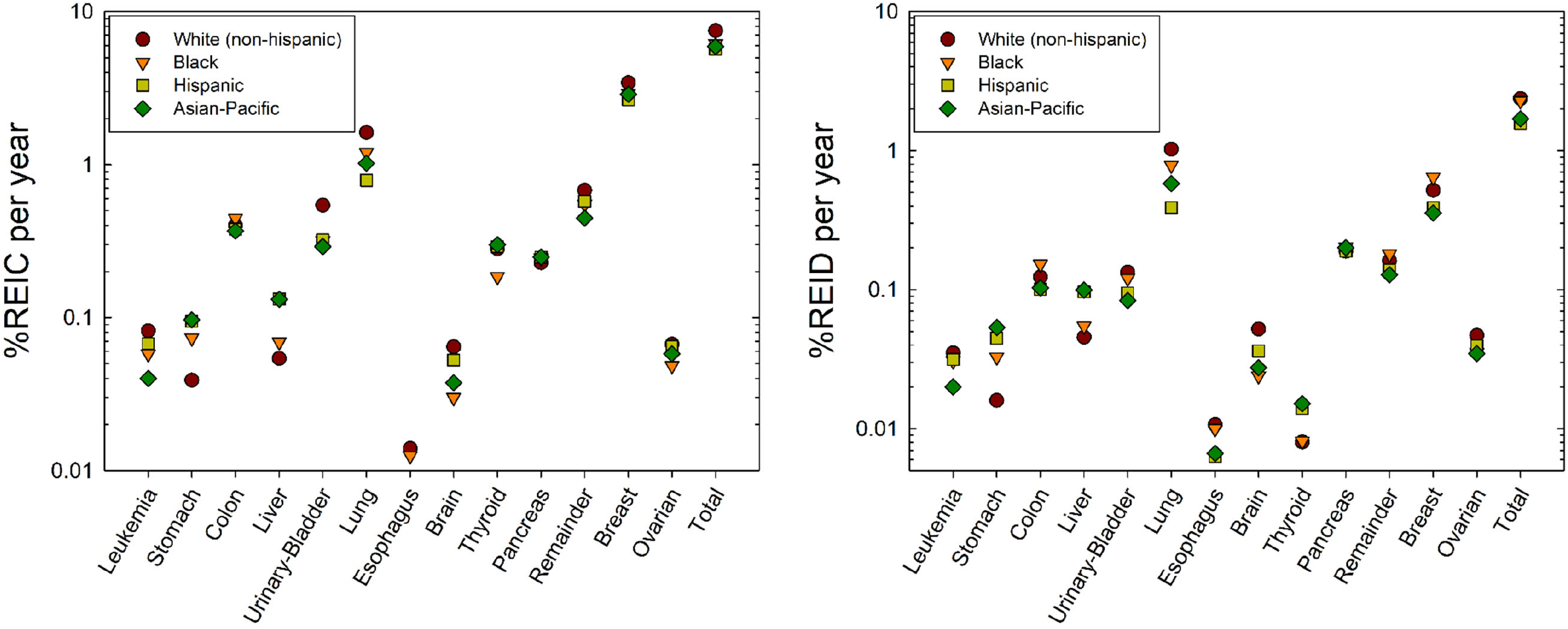
A) Predictions of risk of exposure induced cancer (REIC) for 35 y old U.S. females of different races or ethnic groups for 1-year galactic cosmic ray exposures near solar minimum. B) Predictions of risk of exposure induced death (REID) for 35 y old U.S. females of different races or ethnic groups for 1-year galactic cosmic ray exposures near solar minimum.

**Figure 3.**
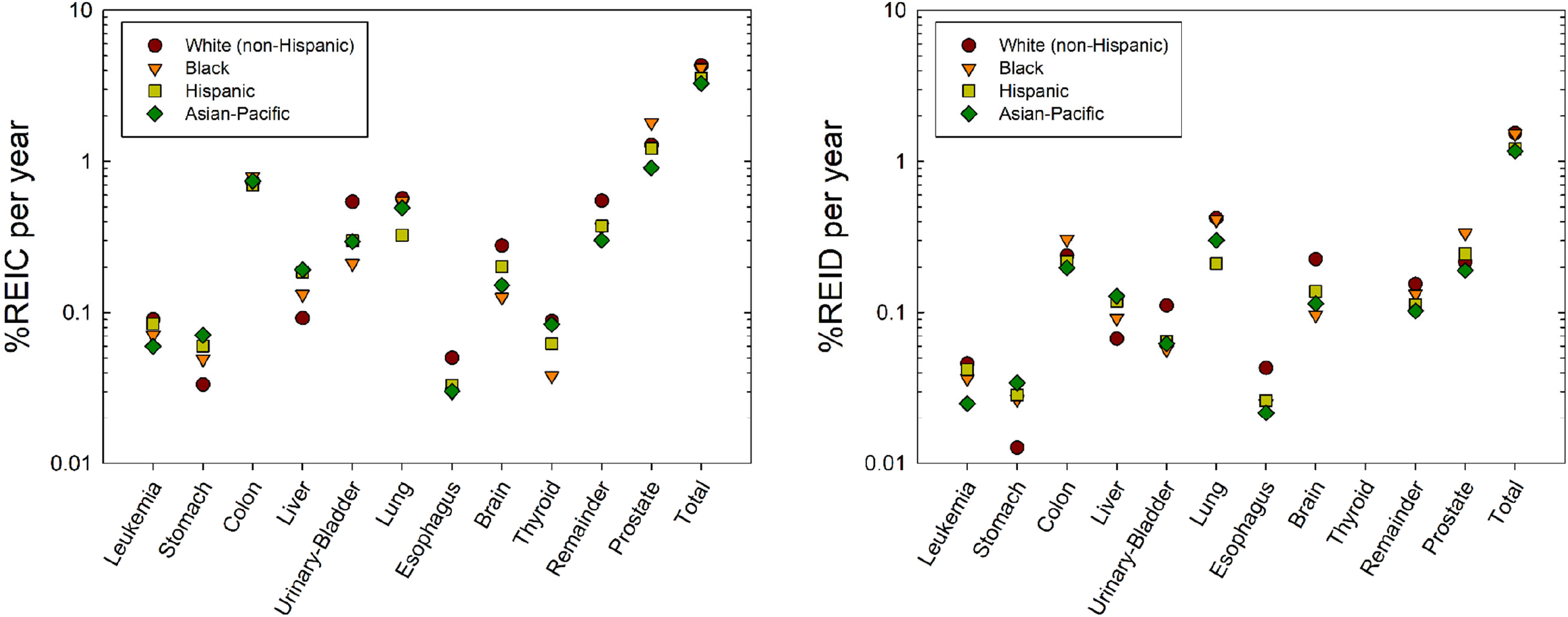
A) Predictions of risk of exposure induced cancer (REIC) for 35 y old U.S. males of different races or ethnic groups for 1-year galactic cosmic ray exposures near solar minimum. B) Predictions of risk of exposure induced death (REID) for 35 y old U.S. males of different races or ethnic groups for 1-year galactic cosmic ray exposures near solar minimum.

**Figure 4**. shows comparison of REIC predictions for females and males of identical race or ethnic group. The relative risks of females to males is to a large extent common to each race or ethnic group, however some differences occur. Females have larger radiation induced stomach cancer risks in all groups except for APIs where females and males have nearly identical risks. The disparity in lung cancer risk with females having much larger than males is reduced to a large extent in APIs.

**Figure 4.**
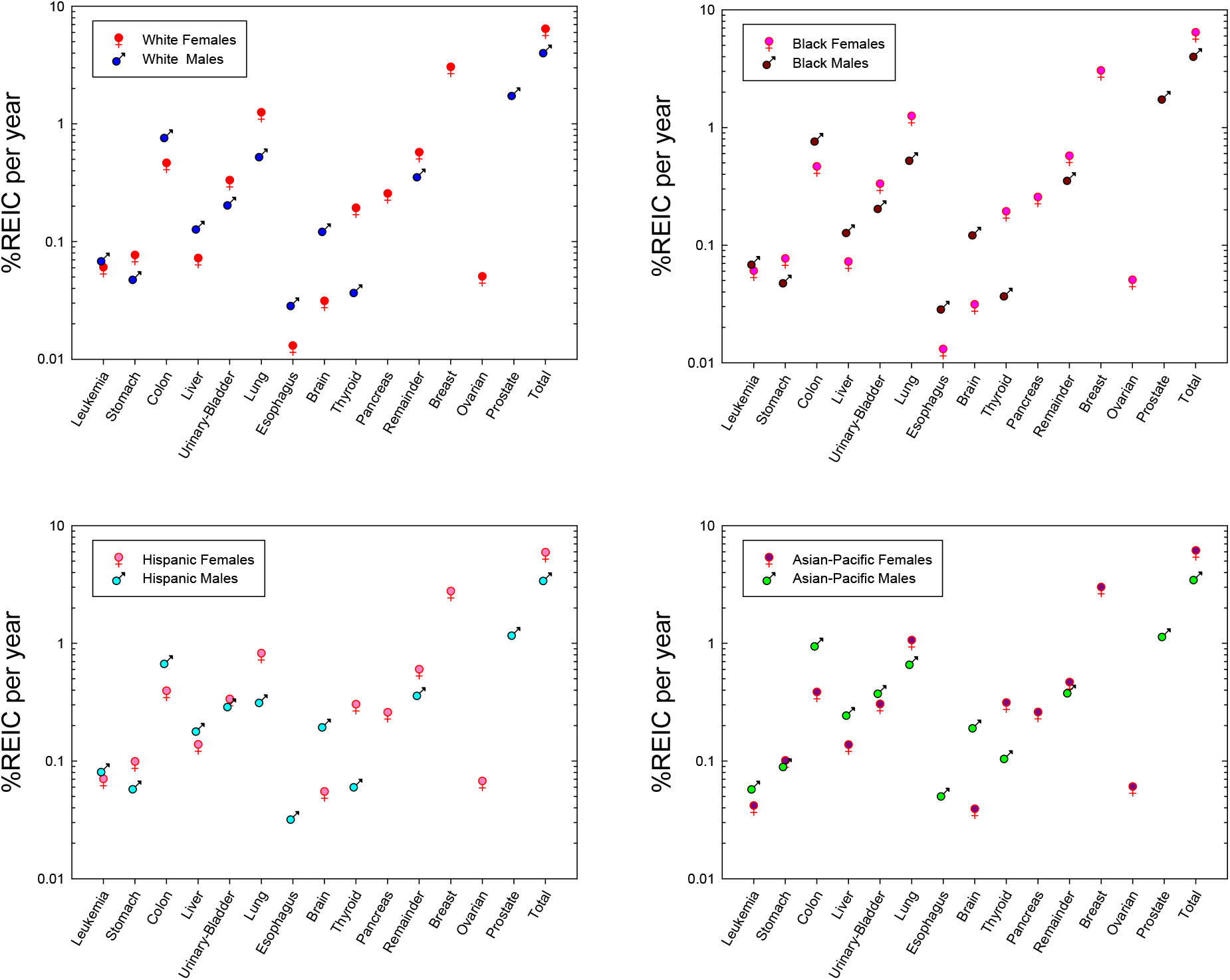
Comparison between female and male risks of exposure induced cancer (REIC) for 35 y old females and males of different races or ethnic groups.

Age is an important factor in the expression of cancer, and the role of the remaining lifespan at older ages due to competing risks reduces radiation risk for increasing ages of exposure. However, it should be noted that the fraction of life remaining compared to the unirradiated for radiation cancer death is nearly constant with age at exposure. **Figure 5** shows predictions of the REIC versus age at exposure from 20 to 60 years of age. Females have the larger risk at all ages at exposure with White females having the largest risk. Females have a larger decline in REIC above age 40 y compared to males due to the large role of declining breast cancer risk with age of exposure. Results using the average US population rates would be slightly reduced compared to predictions for Whites.

**Figure 5.**
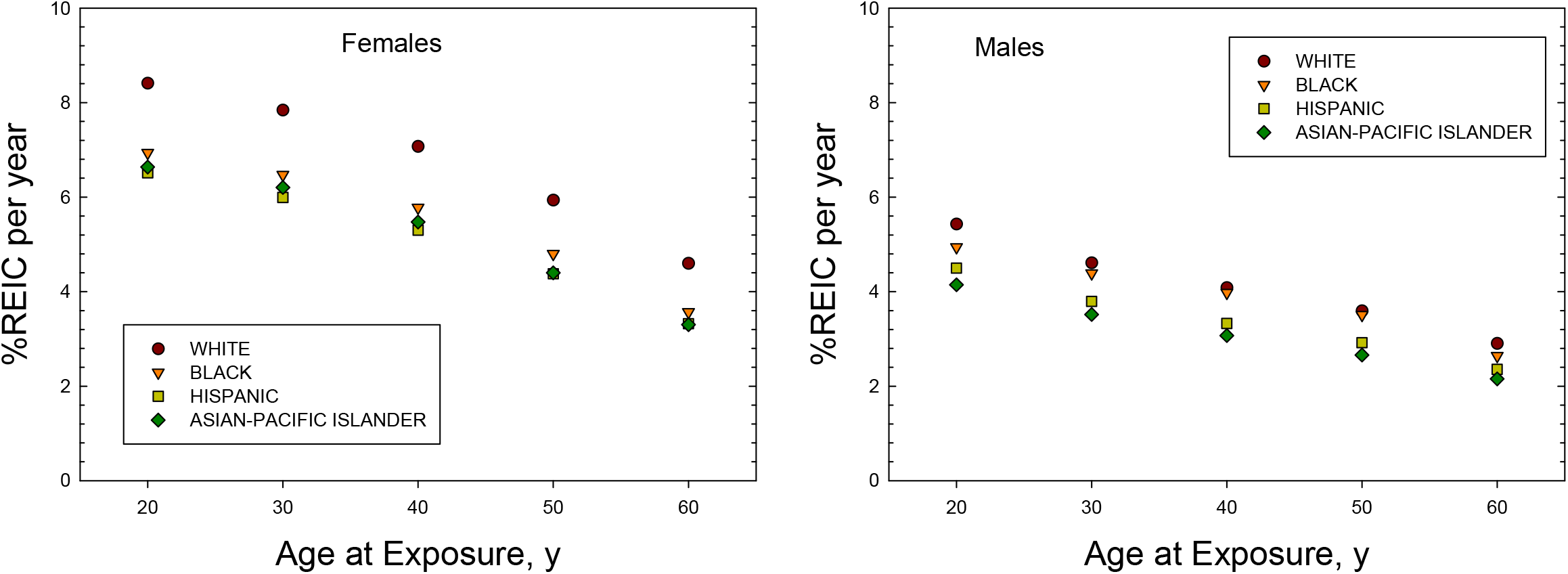
Prediction of risk of exposure induced cancer (REIC) versus age of exposure for females of different races or ethnic groups from 1-year GCR exposures. B) Prediction of risk of exposure induced cancer (REIC) versus age of exposure for males of different races or ethnic groups from 1-year GCR exposures.

We next predicted cancer risks for a references Mars mission of 940 days (400 days transit time and 540 days on the Mars surface) [31]. We considered estimates for a mission near average solar minimum conditions and used the shielding of Mars atmosphere as described previously [32]. Calculations are made in the conventional targeted-effects (TE) model and a model with non-targeted effects (NTE) as described previously [23-26]. Because there are no model factors that describe the interaction of radiation with the underlying causes of different background rates, we report in **Table 4** only the lowest and highest risk populations which are API’s and Whites, respectively. Age at exposure is the most important factor in controlling risk, however the results of **Table 4** suggest differences on the order of 30% and 100% occur between the most sensitive and resistant females and males of a given age. These differences are larger than possibilities for radiation shielding (e.g., aluminum, water, polyethylene or carbon composites) [33]. Non-targeted effects, as described previously [23-26], would lead to large increases in Mars mission risks of a factor of 2 or more.

**Table 4.**
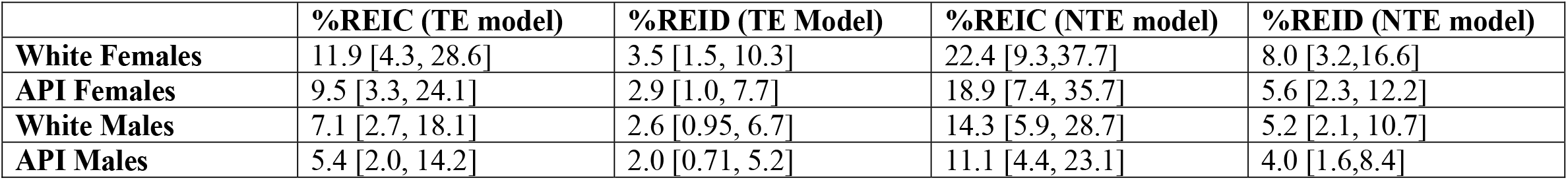
Risk predictions for 35 y old crew persons for 940 d Mars mission. Mission parameters are 400 days total transit and 540 days on surface during average solar minimum conditions with average of 20 g/cm^2^ aluminum shielding. The conventional (targeted effects (TE)) model and model with estimates of non-targeted effect (NTE) as described in the methods section and previously [23-26] are shown. Results for the highest risk group (Whites) and lowest risk group (Asian-Pacific Islanders (API)) are shown.

Our predictions do not account for underlying mechanisms related to genetic, dietary and environmental factors that influence spontaneous cancers [34-36] in relation to how damage induced by protons and heavy ions in space could interact with spontaneous processes impacting cancer risk. However, we suggest that research in this area is clearly warranted. High LET radiation produces not only more complex forms or DNA damage, but also modulates signaling pathways in a distinct manner to other radiation types [8-11], with unknown implications for risks in individuals with distinct racial and ethnic backgrounds.

In conclusion, we find important differences in predictions of space radiation cancer risks for several racial and ethnic groups. These differences suggest inadequacies in risk predictions that consider only the U.S. average population, with possible implications for advising individuals of their potential risks and minimizing risks for specific missions. The science and legal status of genetic based risk evaluations is immature at this time [37]. We suggest that assessing risks based on age, sex and racial and ethnic background should be considered in an operational context for space missions in both the private and government sectors. Interestingly, astronaut selection is based on variety of factors such as requirements for vision, blood pressure, height and weight, and a degree in STEM education discipline [38,39]. Each of these criteria has likley genetic, racial, and environmental contributions. We suggest for high risk missions; similar considerations should be considered in crew selection if the result is a lower mission radiation risk.

## Methods

We briefly summarize recent methods developed to predict the risk of exposure induced cancer (REIC) and risk of exposure induced death (REID) for space missions and associated uncertainty distributions [23-27]. The instantaneous cancer incidence or mortality rates, λ_I_ and λ_M_, respectively, are modeled as functions of the tissue averaged absorbed dose *D*_*T*_, or dose-rate *D*_*Tr*_, gender, age at exposure *a*_*E*_, and attained age *a* or latency *L*, which is the time after exposure *L*=*a-a*_*E*_. The λ_I_ (or λ_M_) is a sum over rates for each tissue that contributes to cancer risk, λ_IT_ (or λ_MT_). These dependencies vary for each cancer type that could be increased by radiation exposure. The REIC is calculated by folding the instantaneous radiation cancer incidence-rate with the probability of surviving to time *t*, which is given by the survival function *S*_*0*_*(t)* for the background population times the probability for radiation cancer death at previous time, summing over one or more space mission exposures, and then integrating over the remainder of a lifetime:

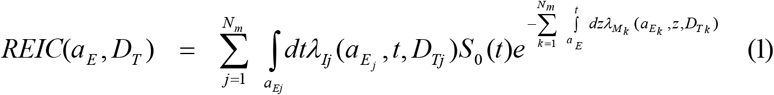

where z is the dummy integration variable. In equation (1), N_m_ is the number of missions (exposures), and for each exposure, j, there is a minimum latency of 5-years for solid cancers, and 2-years for leukemia assumed. Tissue specific REIC estimates are similar to equation (1) using the single term from λ_I_ of interest. The equation for the REID estimate is similar to equation (1) with the incidence rate replaced by the mortality rate (defined below). We terminate the integral in Eq.(1) at the age of 100 years.

The tissue-specific cancer incidence rate for an organ absorbed dose, *D*_*T*_, is written as a RR model after adjustment for radiation quality and dose-rate through introduction a function R_QF_:

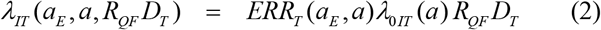

where *λ*_*0IT*_ is the tissue-specific cancer incidence rate in the reference population, and *ERR*_*T*_ the tissue specific excess relative risk per Sievert. Extension of Eq. (2) for a spectrum of particle is described below. The tissue specific rates for cancer mortality *λ*_*MT*_ are modeled following the BEIR VII report [30] whereby the incidence rate of equation (2) is scaled by the age, sex, and tissue specific ratio of rates for mortality to background incidence in the population under study:

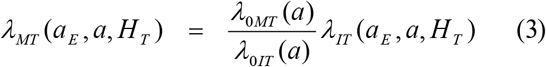

Lifetables from the U.S. Center of Disease Control and Prevention (CDC) for white, black and Hispanic (black and white) male and females are used [4], while the life-table for Asian-Pacific Islander populations are adjusted from the data for the Hispanics for their higher life expectance of ∼2 to 3 years. Total and tissue specific cancer incidence and mortality rates from U.S. SEER are used with data collected from 2014-2018, which provide race, age, and sex specific rates. For cancer incidence we used the SEER delay-adjusted rates, which estimate the impact of delay in reporting of cancer cases [2,3]. In several cases (e.g., liver and brain cancers), the delay-adjusted rates led at older ages (>80 y) to age-specific mortality rates greater than the delay adjusted incidence rate. For these cases we applied a correction to the mortality rates to ensure the mortality rate was 10% larger than the delay adjusted age specific incidence rates.

### ERR Functions

The ERR function was parameterized for various solid cancer to depend on age at exposure, *a*_*E*_, attained aged, *a* using the parametric form [14-22]:

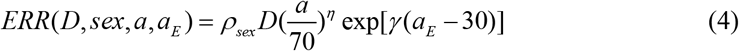

Values for the parameters (ρ,γ, and η)from several reports are shown in **Table 3**. For lung cancer we use the results from a generalized multiplicative model [15], which includes the effects of tobacco usage, however with the number of cigarettes per day set to 0. For leukemia risk excluding CLL uses a dependent on time since exposure in place of age at exposure [13]:

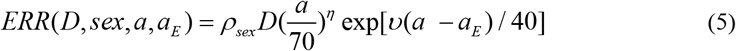

with parameter values also listed in Table 3. For thyroid and remainder (other) cancers we use similar parametrizations from the BEIR VII report [30]. Estimates for uterine cancer were small and not included in the results section.

### Radiation Quality and Dose-Rate Descriptions

The radiation rates in Eq. (2) and (3) are scaled for GCR ions at low dose-rates to gamma-rays using a scaling factor denoted, *R*_*QF*_. The *R*_*QF*_ has two terms that represent the track core and penumbra contributions to ion effects. The parameters of *R*_*QF*_ are estimated from relative biological effectiveness factors (RBE’s) determined from low dose and dose-rate particle data relative to acute γ-ray exposures for doses of about 0.5–3 Gy, which we denote as *RBE*_*γAcute*_ to distinguish from estimates from *RBE*_*max*_ based on less accurate initial slope estimates and a DDREF estimated from Bayesian analysis described previously. The penumbra term contains a dose and dose-rate reduction effectiveness factor (DDREF). The scaling factor is written [23-26]:

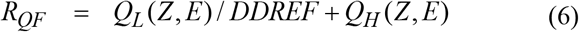

where

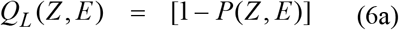

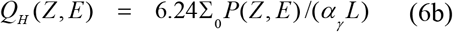

with the parametric function [23-26]

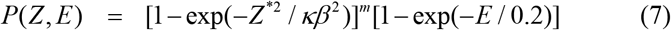

where *E* is the particles kinetic energy per nucleon in units of MeV/u, *L* is the LET, *Z* is the particles charge number, *Z\** the effective charge number, and *β* the particles speed relative the speed of light. The model parameters (Σ_0_/α_γ_, κ and *m*) in Eq.’s (6) and (7) are fit to radiobiology data for tumors in mice or surrogate cancer endpoints as described previously [23-26]. Distinct parameters are used for estimating solid cancer and leukemia risks based on estimates of smaller RBEs for acute myeloid leukemia and thymic lymphoma in mice compared to those found for solid cancers.

The space radiation QF model corresponds to a pseudo-action cross section (biological effectiveness per unit fluence) of the form,

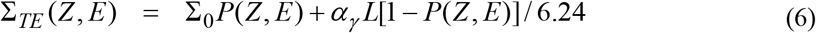

The Σ is denoted as a pseudo-biological action cross section for tumor induction in units of µm^2^ with the designation as “pseudo” given because time-dependent factors have been suppressed, which impact values for the cross-sectional area predicted by fits to the experiments.

### Non-Targeted Effects on QF

In the NTE model we assume the TE contribution is valid with a linear response to the lowest dose or fluence considered, while an additional NTE contribution occurs such a pseudo-action cross section is given by,

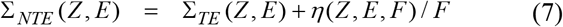

where *F* is the particle fluence (in units of µm^2^) and the η function represents the NTE contribution, which is parameterized as a function of *x=Z*^*\*2*^*/β*^*2*^ or similarly *LET* as:

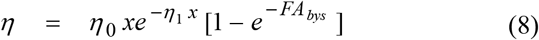

In Eq. (8) the area, *A*_*bys*_, determines the number of bystander cells surrounding a cell traversed directly by a particle that receives an oncogenic signal. The RBE (or QF) is related to the cross section by *RBE = 6*.*24 Σ/(LET α*_*γ*_) where α_γ_ is the gamma-ray linear slope coefficient. Therefore, only the ratio of parameters η_0_/α_γ_ is needed for risk estimates.

### Space Radiation Organ Exposures

GCR exposures include primary and secondary H, He and HZE particles, and secondary neutrons, mesons, electrons, and γ-rays over a wide energy range. We used the HZE particle transport computer code (HZETRN) with quantum fragmentation model nuclear interaction cross sections and Badhwar–O’Neill GCR environmental model to estimate particle energy spectra for particle type *j, φ*_*j*_*(Z,E)* as described previously [23,32]. GCR organ dose equivalent show little variation from 10 to 50 g/cm^2^ of shielding [33], and we use 20 g/cm^2^ for calculations, which is a typical average shielding amount.

For the TE model, a mixed-field pseudo-action cross section is formed by weighting the particle flux spectra, *φ*_*j*_*(E*) for particle species, *j*, contributing to GCR exposure evaluated with the HZETRN code with the pseudo-biological action cross section for mono-energetic particles and summing over all particles and kinetic energies:

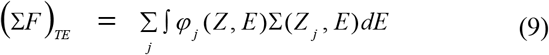

Equations for the mixed-field pseudo-action cross section in the NTE model as folded with particle specific energy spectra as:

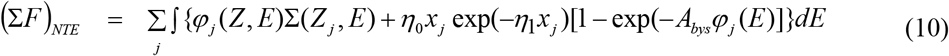

Further details on uncertainty analysis of model components are described in previous reports [23-27]. We note that estimates of uncertainties in particle spectra and organ doses, dose-rate modifiers, and radiation quality function parameters are included in our approach. PDF’s for each component are formulated and Monte-Carlo sampling performed to propagate the uncertainty over each factor to obtain an estimate of the overall uncertainty.

## Data Availability

All data is available in literature or SEER and CDC WEB.

## References

1. Von der Dunk, F. G. Space tourism, private spaceflight and the law: Key aspects”. Space Policy 27, 146–152 (2011).

2. SEER Explorer: An interactive website for SEER cancer statistics [Internet]. Surveillance Research Program, National Cancer Institute. [Cited 2021 April 15]. Available from https://seer.cancer.gov/explorer/.

3. Midthune, D.N., Fay, M.P., Clegg, L.X., Feuer, EJ. Modeling reporting delays and reporting corrections in cancer registry data. J Am Stat Assoc 100, 61–70 (2005).

4. National Vital Statistics Report, U.S. Life Table 2018 Vol 70, No 1 (2021)

5. Barcellos-Hoff M.H., et al. The evolution of the cancer niche during multistage carcinogenesis. Nature Rev Cancer 13, 511–518 (2013).

6. Hanahan, D., Weinberg R.A. Hallmarks of cancer: the next generation. Cell 144, 646–675 (2011).

7. Fry RJM. Experimental radiation carcinogenesis: what have we learned? Radiat Res 87, 224–239 (1981).

8. Cucinotta, F.A., Durante, M. Cancer risk from exposure to galactic cosmic rays: implications for space exploration by human beings. Lancet Oncol 7, 431–435 (2006).

9. Sridharan D.M., et al. Understanding cancer development processes following HZE particle exposure: roles of ROS, DNA damage repair, and inflammation. Radiat Res 183, 1–26 (2015).

10. Omene C, et al. Aggressive mammary cancers lacking lymphocytic infiltration arise in irradiation mice and can be prevented by dietary intervention. Cancer Immun Res 8, 217–229, (2020).

11. Kadhim, M. et al. Non-targeted effects of ionizing radiation-implications for low dose risk. Mut Res 752, 84–98 (2013).

12. Storer JB, Mitchell TJ, Fry RJM. Extrapolation of the relative risk of radiogenic neoplasms across mouse strains and to man. Radiat Res 114, 331–353 (1988).

13. Hsu, W., et al. The incidence of leukemia, lymphoma, and multiple myeloma among atomic bomb survivors: 1950-2001. Radiat Res 179, 361–382 (2013).

14. Grant, E.J., et al. Solid cancer incidence among the Life-span study of atomic-bomb survivors: 1958-2009. Radiat Res 187, 513–537 (2017).

15. Cahoon, E.K. et al. Lung, laryngeal and other respiratory cancer incidence among Japanese atomic bomb survivors: an updated analysis from 1958 through 2009. Radiat Res 187, 538–548 (2017).

16. Brenner, A.V., et al. Incidence of breast cancer in the Life Span Study of the atomic bomb survivors: 1958-2009. Radiat Res 190, 433–444 (2018).

17. Sadakane, A., et al. Radiation and risk of liver, biliary tracts, and pancreatic cancers in the atomic bomb survivors in Hiroshima and Nagasaki: 1958-2009. Radiat Res 192, 299–310 (2019).

18. Sakata, R., et al. Radiation-related risk of cancers of the upper digestive tract among Japanese atomic bomb survivors. Radiat Res 192, 331–344 (2019).

19. Brenner, A.V. et al. Radiation risk of central nervous system tumors in the Life Space Study of atomic bomb survivors, 1958-2009. Eur J Epidem 35, 591–600 (2020).

20. Sugiyami, H., et al. Radiation risk of incident colorectal cancer by anatomic site among atomic bomb survivors: 1958-2009. Int J Cancer 146, 635–645 (2019).

21. Mabuchi, K., Risk of prostate cancer incidence among atomic bomb survivors: 1958-2009. Radiat Res 195, 66–76 (2021).

22. Utada, M., Radiation risk of ovarian cancer in atomic bomb survivors: 1958-2009. Radiat Res 195, 60–65 (2021).

23. Cucinotta, F.A., To, K., Cacao, E. Predictions of space radiation fatality risk for exploration missions. Life Sci Space Res 13, 1–11 (2017).

24. Cucinotta, F.A., Cacao, E. Non-targeted effects models predict significantly higher mars mission cancer risk than targeted effects model. Scientific Rep 7, 1832 (2017).

25. Cucinotta, F.A., Cacao, E., Kim, M.Y., Saganti, P.B. Cancer and circulatory disease risks for a human mission to Mars: private mission considerations. Acta Astronautica 166, 529–536 (2020).

26. Cucinotta, F.A., Cacao, E., Kim, M.Y., Saganti, P.B. Benchmarking risk predictions and uncertainties in the NSCR model of GCR cancer risks with revised low LET risk coefficients. Life Sci Space Res 27, 64–73 (2020).

27. Cucinotta, F.A. Space radiation risks for astronauts on multiple International Space Station missions. PLoS ONE 9(4), e96099 (2014).

28. Acciai, F., Noah, A.J., Firebaugh, G. Pinpointing the sources of the Asian mortality advantage in the United States. J Epidemiol Community Health 69, 1006–1011 (2015).

29. Torre, L.A., et al. Cancer statistics for Asian Americans, Native Hawaiians, and Pacific Islanders, 2015: convergence of incidence between males and females. CA Cancer J Clin 66, 182–202 (2016).

30. Beir VII. Health risks from exposure to low levels of ionizing radiation. National Academy of Sciences Committee on the Biological Effects of Radiation. Washington DC: National Academy of Sciences Press (2006).

31. National Aeronautics and Space Administration. Human exploration of mars: design reference architecture 5.0. NASA SP 2009-502, Washington DC (2009).

32. Kim, M.Y., et al. Comparison of Martian surface ionizing radiation measurements from MSL-RAD with Badhwar-O’Neill 2011/HZETRN model calculations. J Geophys Res 119, 1311–1321 (2014).

33. Cucinotta, F.A., Kim, M.Y., Chappell, L.J. Evaluating shielding approaches to reduce space radiation cancer risks. NASA TM- 2012–217631 (2012).

34. Mitchell, K.A., et al. Comparative transcriptome profiling reveals coding and noncoding RNA differences in NSCLC from African Americans and European Americans. Clin Cancer Res 23, 7412–7425 (2017).

35. Kakarla, M., et al. Race as a contributor to stromal modulation of tumor progression. Cancers (Basel) 13, 2656 (2021).

36. Song, M., et al. Racial differences in genome-wide methylation profiling and gene expression in breast tissues from healthy women. Epigenetics 10, 1177–1187 (2015).

37. National Council of Radiation Protection and Measurements Report No 167. Potential impact of individual genetic susceptibility and previous radiation exposure on radiation risk for astronauts. NCRP Bethesda Md (2010).

38. NASA, Astronaut selection and training. https://www.nasa.gov/centers/johnson/pdf/606877main_FS-2011-11-057-JSC-astro_trng.pdf (2011).

39. ESA. Astronaut selection 2021-22 FAQs. https://www.esa.int/About_Us/Careers_at_ESA/ESA_Astronaut_Selection/Astronaut_selection_2021-22_FAQs (2021).

